# Differential transcriptomic impacts of breastmilk compared to formula milk

**DOI:** 10.1101/2025.05.29.25328551

**Authors:** Duan Ni, Ralph Nanan

**Affiliations:** Sydney Medical School Nepean, The University of Sydney, Sydney, NSW, Australia; Charles Perkins Centre, The University of Sydney, Sydney, NSW, Australia; Nepean Hospital, Nepean Blue Mountains Local Health District, Sydney, NSW, Australia

**Keywords:** Breastmilk, formula milk, breastfeeding, formula feeding, metabolic health, intestinal gluconeogenesis, infants

## Abstract

**Background:** Breastmilk confers numerous benefits to infants relative to formula milk, such as promoting metabolic health and supporting immune homeostasis. However, in-depth mechanistic insights comparing the influence of breastmilk versus formula milk across different compartments are lacking.

**Methods:** Several datasets interrogating the effects of breastmilk versus formula milk were curated, including transcriptomes of infant intestinal organoids, intestinal gene expression microarray, and single cell RNA-seq data for infant peripheral blood mononuclear cells (PBMCs). Comparative analyses were performed, with a specific focus on differences at a signal pathway level.

**Results:** Comparative analyses revealed that intestinal exposure to breastmilk was linked to elevated gluconeogenesis, as well as reprogramming of other metabolic, nutrient-sensing and immune-related pathways. Contrarily, in PBMCs, formula milk feeding was linked to upregulation of apoptosis-related pathways across all PBMC subsets.

**Conclusions:** Breastmilk reprograms the transcriptomic landscapes of infant intestine, particularly promoting intestinal gluconeogenesis. This might explain its metabolic advantages via modulating metabolic homeostasis. Different effects were found in PBMCs, where formula milk is linked to enhanced apoptotic signalling in the infant’s developing immune cell subsets.

## Background

Breastmilk confers multifaceted benefits to infants. Particularly, breastmilk-fed infants exhibit better metabolic health, with reduced risk of obesity, diabetes and other metabolic syndrome later in life [1–4]. Previously, studies addressing these benefits are mostly observational. Our recent microarray analyses have shown that breastfeeding is linked to elevated intestinal gluconeogenesis in infants across multiple mammalian species including humans [5]. Increased intestinal gluconeogenesis is likely to lead to superior metabolic homeostasis of breastmilk-fed infants [6]. In fetal intestinal organoids, breastmilk has recently been shown to support their growth, differentiation and homeostatic cytokine production [7].

Benefits from breastmilk extend beyond the metabolic system. For example, breastfeeding is known to modulate the developing immune system in early life [8–10] and might contribute to protection against the development of allergic diseases [11–13]. This is supported by a recent study reporting that peripheral blood mononuclear cells (PBMCs) from breastfed infants downregulated allergy-related interleukin (IL)-4 and IL-13 signalling pathways [14]. There is now a growing body of evidence showing that immune cells and functions are tightly regulated by their metabolisms [15–17]. In this context, whether breastmilk might rewire the immunometabolic landscape of infants and further modify their immune functions warrants more thorough interrogation.

Here, curating published transcriptomic datasets of fetal intestine and intestinal organoid and a single cell RNA-seq (scRNA-seq) dataset of infant PBMCs, we comprehensively compared the impact of breastmilk versus formula milk on the infants’ transcriptomic landscapes, with a particular focus on their immunometabolism. We corroborated that breastmilk led to increased intestinal gluconeogenesis and extensively reprogramed the fetal intestinal transcriptome. In contrast, PBMCs from infants fed on formular milk displayed an increase in apoptosis-related signalling. These observations highlight the differential impacts from breastmilk versus formula milk on infants in different compartments.

## Methods

RNA-seq [7], microarray [18], and scRNA-seq dataset [14] were retrieved from their original publications and downloaded from Gene Expression Omnibus (GEO) respectively (GEO ID: GSE253501, GSE31075, GSE296678). For RNA-seq dataset, organoid samples cultured in growth media supplemented with parental milk (breastmilk) and extensively hydrolyzed formula (formula milk) were selected for analysis. Raw data from RNA-seq and microarray experiments were first normalized with *DESeq2* package [19] before analysis with Gene Set Enrichment Analysis (GSEA) software [20]. For scRNA-seq dataset, it was analyzed with *Seurat* package [21]. Different individual samples were integrated with *LIGER* package [22]. GSEA was run using *fgsea* package.

## Results

We first leveraged a transcriptomic dataset (GEO ID: GSE253501) from a study where fetal small intestine organoids were cultured for 5 days in growth media supplemented with either human breastmilk (BM) or extensively hydrolyzed formula (HF) (Figure 1a) [7]. This represents a unique opportunity to explore the direct impacts of breastmilk versus formula milk on fetal intestinal biology.

**Figure 1.**
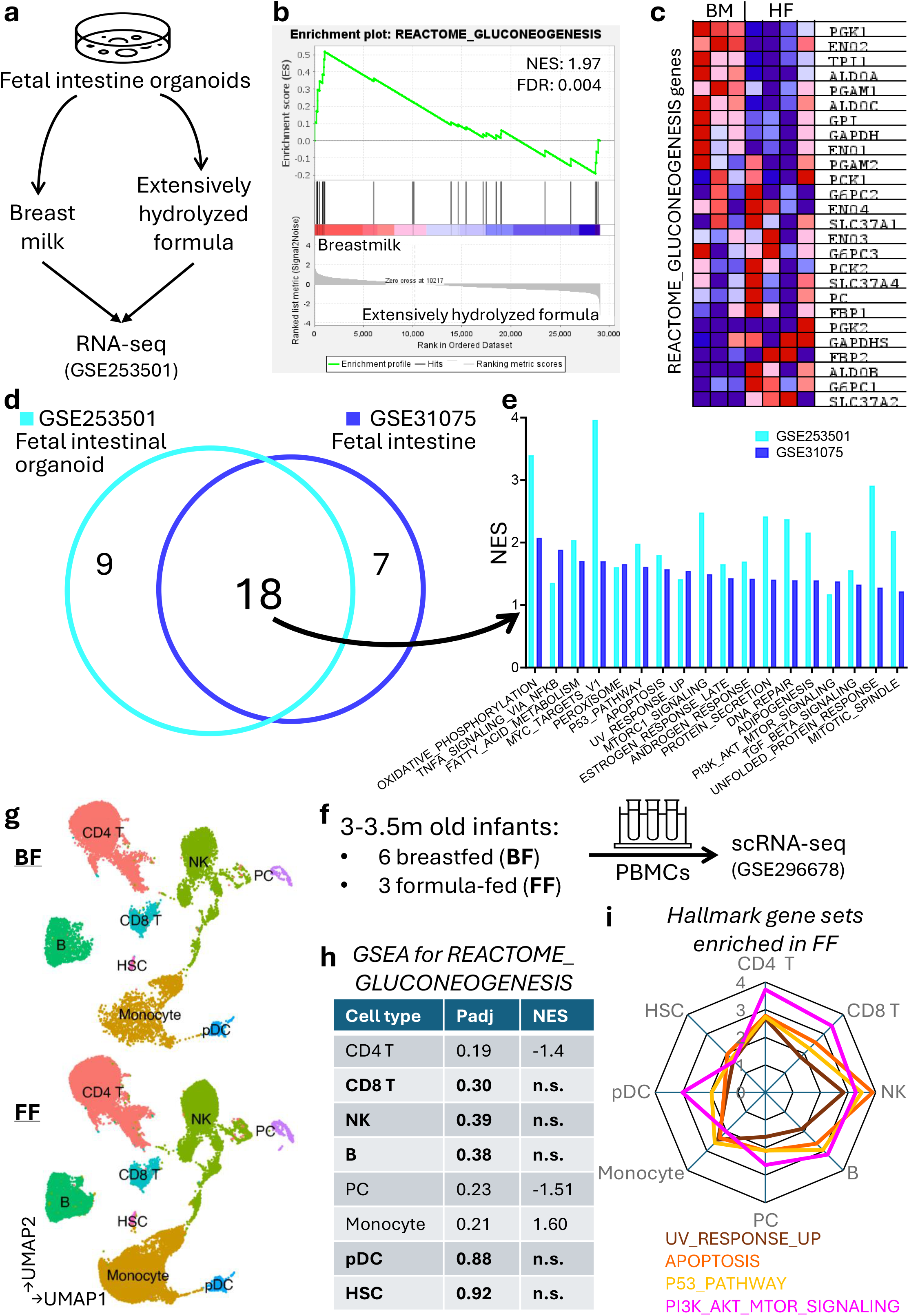
**a**. Overview of fetal intestine organoid data. RNA-seq data was obtained from GSE253501, where fetal small intestine organoids were cultured for 5 days in growth media supplemented with parental milk (breastmilk) or extensively hydrolyzed formula milk (formula milk). Organoids were then subject to RNA-seq analysis. **b**. Gene Set Enrichment Analysis (GSEA) showed that fetal intestine organoids treated with breastmilk were enriched in “REACTOME_GLUCONEOGENESIS” gene set relative to the ones treated with extensively hydrolyzed formula. **c**. Heatmap for expression of genes within “REACTOME_GLUCONEOGENESIS” gene set in fetal intestine organoids treated with breastmilk (BM) or extensively hydrolyzed formula (HF), with red indicating high expression and blue reflecting low expression. **d**. Comparison of gene sets enriched in fetal intestine organoids treated with breastmilk (cyan, analysis based on GSE253501, 27 enriched) and in fetal intestines from infants fed on breastmilk (blue, analysis based on GSE31075, 25 enriched) based on GSEA with 50 Hallmark Gene Sets. **e**. Details of the 18 gene sets consistently enriched in fetal intestine organoid treated with breastmilk and in fetal intestines from infants fed on breastmilk based on analysis in **d. f**. Overview of peripheral blood mononuclear cell (PBMC) single cell RNA-seq (scRNA-seq) data. scRNA-seq data was retrieved from GSE296678, where PBMCs were isolated from 3-3.5 months old breastfed (6, BF) or formula-fed (3, FF) infants and were then analyzed by scRNA-seq. **g**. Uniform manifold approximation and projection (UMAP) plots visualizing the BF (upper) and FF (lower) infant PBMC cellular compositions by scRNAseq (CD4^+^ T cell (CD4 T), CD8^+^ T cell (CD8 T), natural killer (NK) cell, B cell, plasma cell (PC), monocyte, plasmacytoid dendritic cell (pDC) and hematopoietic stem cell (HSC)). **h**. Summary of GSEA results for “REACTOME_GLUCONEOGENESIS” gene set comparing different BF vs FF PBMC subsets. Subsets in bold signified no significant difference was found. **i**. Radar plot visualizing the normalized enrichment scores for “HALLMARK_PI3K_AKT_MTOR_SIGNALING” (magenta), “HALLMARK_P53_PATHWAY” (yellow), “HALLMARK_APOPTOSIS_PATHWAY” (orange)

GSEA found that organoids exposed to breastmilk were significantly enriched in “REACTOME_GLUCONEOGENESIS” pathway (Figure 1b-c), aligning with our previous findings that breastmilk-fed infants exhibited upregulated intestinal gluconeogenesis [5]. To more comprehensively interrogate the impacts from breastmilk on intestinal biology, we performed GSEA for these two datasets of infant intestine organoids (GEO ID: GSE253501) [7] and infant intestine (GEO ID: GSE31075) [18] with Hallmark Gene Sets, which covered 50 representative pathways related to major biological statuses and processes. 27 (GSE253501) and 25 (GSE31075) Hallmark Gene Sets were enriched in organoids cultured with breastmilk and breastmilk-fed infants’ intestines respectively (Figure 1d). Among them, breastmilk-related increases in 18 gene sets were consistently detected in both contexts (Figure 1e), including metabolic processes like oxidative phosphorylation and fatty acid metabolism, nutrient-sensing pathways like mTOR signals, and immune-related signalling involving TNF-α and TGF-β. No consistent change was found when comparing fetal intestinal organoid exposed to formula and formula milk-fed infants. Together, these data confirmed our previous report that breastfeeding was linked to elevated intestinal gluconeogenesis [5] and corroborated the substantial shift of intestinal transcriptomic landscapes linked to breastmilk.

We next ask whether these breastmilk-mediated transcriptomic reprogramming would extend to other compartments. A scRNA-seq data set (GEO ID: GSE296678) was retrieved from recent work by Salinas *et al*. [14], which profiled the PBMCs from 3-3.5 months old infants fed on breastmilk (BF, n=6) versus formula milk (FF, n=3) (Figure 1f). Among 8 populations identified (CD4^+^ T cell, CD8^+^ T cell, natural killer (NK) cell, B cell, plasma cell (PC), monocyte, plasmacytoid dendritic cell (pDC) and hematopoietic stem cell (HSC)), FF PBMCs had higher proportion of monocytes (Figure 1g), aligning with their findings [14]. Surprisingly, GSEA found limited differences for “REACTOME_GLUCONEOGENESIS” pathway comparing BF and FF. In BF PBMCs, gluconeogenesis signal was higher in monocytes but lower in CD4^+^ T cell and PC and was insignificantly altered in other subsets (Figure 1h). GSEA based on Hallmark Gene Sets were run for all 8 PBMC subsets. No pathway was consistently increased in BF samples. Contrarily, in FF PBMCs, all populations were constantly enriched in gene sets “HALLMARK_PI3K_AKT_MTOR_SIGNALING”, “HALLMARK_P53_PATHWAY”, “HALLMARK_APOPTOSIS_PATHWAY” and “HALLMARK_UV_RESPONSE_UP” (Figure 1i).

## Discussions

Here, we present to the best of our knowledge the first comprehensive study comparing the effect of breastmilk with formula milk across different compartments. Using a fetal intestinal organoid dataset, we validated our prior finding that breastfeeding is associated with increased intestinal gluconeogenesis [5]. We also uncovered other transcriptomic changes upon exposure to breastmilk, including shifts in metabolic and nutrient-sensing pathways and alterations in immune signalling. Contrarily, in infants’ PBMCs, gluconeogenesis pathway was inconsistently impacted by breastmilk. Formula milk feeding was linked to upregulation of PI3K-Akt-mTOR and p53 signals and apoptosis pathway. Collectively, these data suggested differential compartmentalized impacts of breastmilk and formula milk.

We previously found that across multiple mammalian species including humans, breastfeeding is linked to upregulated intestinal gluconeogenesis in infants [5], but the detailed causality remained elusive. Our results here based on fetal intestinal organoid demonstrated the direct effect of breastmilk on fetal intestinal gluconeogenesis. Along this stream, the organoid system might also be used to examine different components of breastmilk to unravel the detailed triggers for the upregulated intestinal gluconeogenesis. Multiple other factors also regulate intestinal gluconeogenesis *in vivo* [6]. Our previous study suggested that the effects of breastmilk was unlikely to be mediated via the gut microbiota [5, 23] but impacts on other aspects like hormonal or neural signals warrant further interrogations. Moreover, whether the gluconeogenic effects from breastmilk might extend to other organs such as liver and kidney also remains to be elucidated, as here we showed limited change of gluconeogenesis in circulating immune cells. Since intestinal gluconeogenesis is critical for glucose homeostasis maintenance [6, 23], our findings here might provide some direct evidence for the metabolic benefits of breastmilk and breastfeeding.

In addition to gluconeogenesis, metabolic and nutrient-sensing signalling like fatty acid metabolism and mTOR pathway exhibited upregulation linked to breastmilk in infant intestine as well. Whether these metabolic alterations might contribute to the immune changes we observed like higher TNF-α and TGF-β signalling in the fetal intestine require further interrogations.

In PBMCs, breastmilk seemed to exert less effects. In contrast, formula milk feeding was linked to an expanded monocyte population, which requires further validation with larger cohorts. Across all developing PBMC subsets, formula milk was associated with elevations in apoptosis-related signals such as the “P53_PATHWAY” and “APOPTOSIS_PATHWAY” gene sets. This might imply deviation from normal immune development seen in breastmilk-fed infants. These changes underpin previous reports that formula feeding negatively influenced infant immunity relative to breastmilk feeding.

## Conclusions

Collectively, we here present the first comprehensive characterization of the differential effects from breastmilk and formula milk on infant intestine and PBMCs. These observations provide some mechanistic insights into the immune and metabolic benefits of breastmilk and breastfeeding compared to formula feeding.and “HALLMARK_UV_RESPONSE_UP” (brown) in FF PBMC subsets relative to their BF counterparts. (NES: normalized enrichment score, FDR: false discovery rate, n.s.: not significant)

## List of abbreviations

BF: fed on breastmilk
BM: breastmilk
Extensively hydrolyzed formula: HF
FF: fed on formula milk
GEO: Gene Expression Omnibus
GSEA: Gene Set Enrichment Analysis
HSC: hematopoietic stem cell
Interleukin: interleukin
NK cell: natural killer cell
PBMCs: peripheral blood mononuclear cells
PC: plasma cell
pDC: plasmacytoid dendritic cell
scRNA-seq: single cell RNA-seq

## Declarations

### Ethics approval and consent to participate

Not applicable

### Consent for publication

Not applicable

### Availability of data and materials

Data is available as Gene Expression Omnibus ID: GSE253501, GSE31075, GSE296678.

### Authors’ contributions

*Concept and design:* D.N., and R.N..

*Acquisition, analysis, and interpretation of data:* D.N., and R.N..

*Drafting of the manuscript:* D.N., and RN..

*Critical revision of the manuscript for important intellectual content:* All authors.

All authors read and approved the final manuscript.

